# COVID-19 Infections Following Physical School Reopening

**DOI:** 10.1101/2020.10.24.20218321

**Authors:** Oren Miron, Kun-Hsing Yu, Rachel Wilf-Miron, Isaac S. Kohane, Nadav Davidovitch

**Affiliations:** Department of Health Systems Management, Ben-Gurion University of the Negev, Beer Sheva, Israel; Department of Biomedical Informatics, Harvard Medical School, Boston, Massachusetts; Faculty of Medicine, Tel Aviv University, Tel Aviv, Israel

## Abstract

**Background:** School reopened in August-September 2020 and their effect on COVID-19 infections is unclear.

**Methods:** We examined Coronavirus Disease-19 incidence following school reopening in Florida.

**Results:** We found that counties teaching physically had 1.2-fold incidence increase in elementary schools and 1.3-fold increase in high schools, while counties teaching remotely had no increase.

**Conclusions:** Our results suggest that counties teaching physically could consider teaching remotely, especially in high school, until it is safe to teach physically.

**What was known:** Schools reopened in August-September 2020, with some teaching remotely, since the effect of physical reopening on COVID-19 infections is unclear.

**What we added:** counties teaching physically had 1.2-fold incidence increase in elementary schools and 1.3-fold increase in high schools. This suggests that counties teaching physically could consider teaching remotely instead, especially at the high school level.

## Introduction

The United States closed most of its schools in March-April 2020, which was associated with a later reduction in coronavirus disease 2019 (COVID-19) incidence.^1^ In August-September 2020, elementary schools and high schools have reopened, with many schools teaching remotely until they estimate they can safely teach physically.

Their calculation is based on the current understanding of the tradeoffs of lower COVID-19 incidence in these age groups,^2^ the educational and social costs of school closure versus the risk of spreading the virus to older age groups.^3^ We seek here to quantify the physical reopening risk by examining these associations in a single state.

## Methods

We searched for a state with both physical learning and remote learning, and that has a COVID-19 database that allows calculation of the exact COVID-19 rate at the county level of ages 6-13 years (elementary school) and 14-17 years (high school). The Florida dataset matched these criteria.^4^ We extracted the daily incidence of cases by county, and matched it with each county’s date of school reopening (day 0).^5^ We examined the COVID-19 case rate with a 7-day moving average.

We aggregated the rates by counties with physical learning and counties with remote learning (the latter was only allowed in Broward, Miami-Dade and Palm Beach). We calculated time points at which the trends changed, in the 10 days before reopening and the 20 days from reopening. The trend analysis used the JoinPoint Regression Program (National Cancer Institute). Lastly, we tested for each age group if the slopes from school reopening to day 20 are significantly different between counties teaching physically and remotely. The statistical tests were 2-tailed with significance at under 0.05.

## Results

In counties teaching physically, at ages 6-13 years the incidence decreased from day −10 to day 4 (−0.5% daily-change, 95%-CI −0.9% to −0.1%), followed by an increase from day 4 to day 20 (0.8% daily-change, 95%-CI 0.5% to 1.1%). On day 4, the incidence was 11 per 100,000 (95%-CI 9.9 to 12), and on day 20 it increased to 12.8 (95%-CI 11.7 to 13.9, 1.2-fold; figure 1).

**Figure 1.**
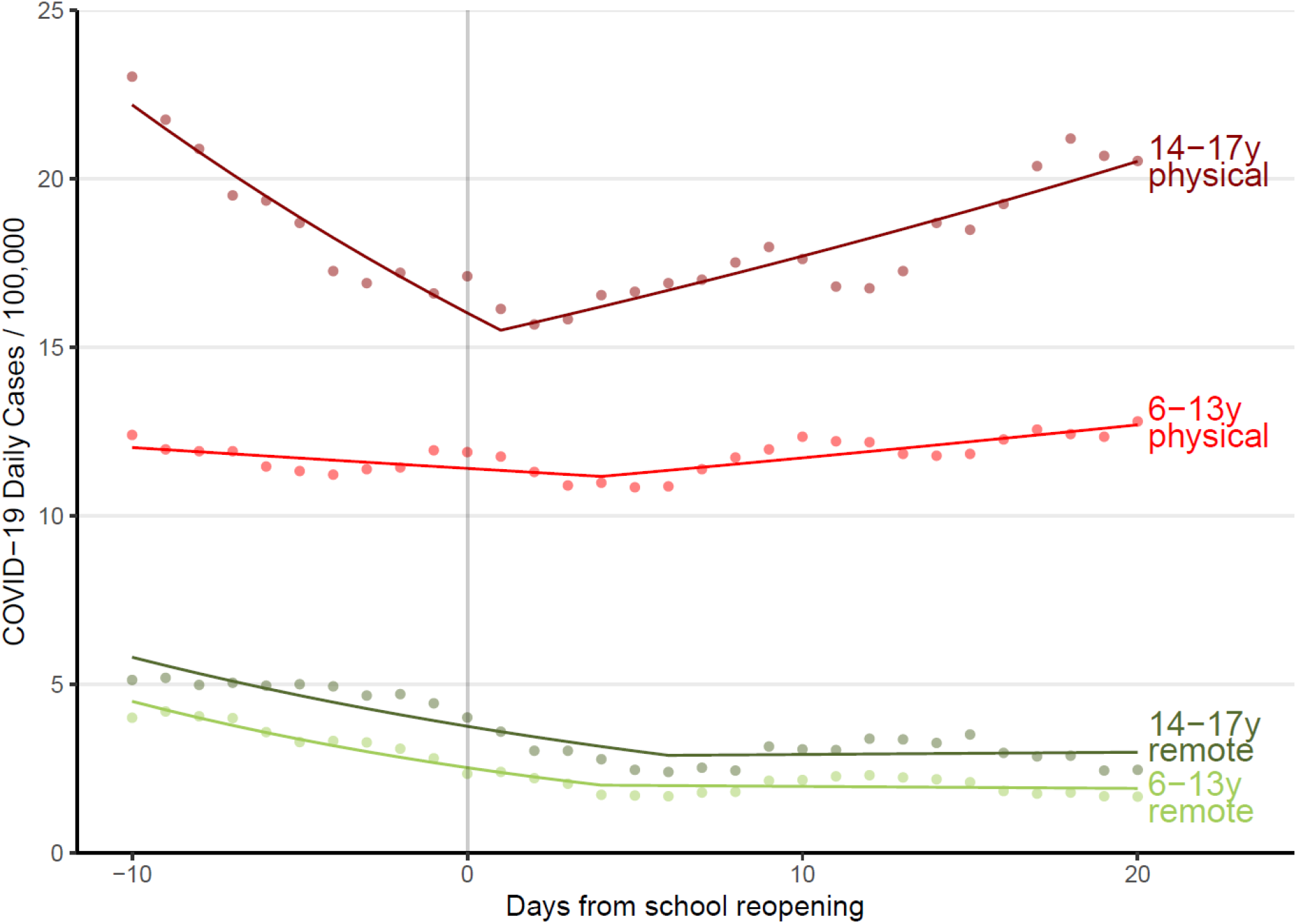
COVID-19 infection rate by days from reopening by age. Caption: Y indicates COVID-19 daily infections per 100,000 capita, and X indicates days from reopening. Dots indicate the observed data points, while the segments of trend lines indicate the JoinPoint regression. Ages 6-13 years in counties with physical learning are in red, and ages 14-17 years in counties with physical learning are in dark red. Ages 6-13 years in counties with remote learning are in green, and ages 14-17 years in counties with remote learning are in dark green.

In counties teaching physically, at age 14-17 years the incidence decreased from day −10 to day 1 (−3.2% daily-change, 95%-CI −3.9% to −2.5%), followed by an increase until day 20 (1.4% daily-change, 95%-CI 1.1% to 1.8%). On day 1, the incidence was 16.1 (95%-CI 14.4 to 17.9), and on day 20 it increased to 20.5 (95%-CI 18.5 to 22.5, 1.3-fold).

In counties teaching remotely, at ages 6-13 years the incidence decreased from day −10 to day 4 (−5.6% daily-change, 95%-CI −6.7% to −4.5%), followed by no significant trend. The slope from day 0 was significantly different than the slope of counties teaching physically at ages 6-13 years (T=5.0, p<0.05).

In counties teaching remotely, at age 14-17 years the incidence decreased from day −10 to day 6 (−4.3% daily-change, 95%-CI −5.4% to −3.1%), followed by no significant trend. The slope from day 0 was significantly different than the slope of counties teaching physically at ages 6-13 years (T=7.6, p<0.05).

## Discussion

Our analysis shows that physical reopening was followed by increased COVID-19 incidence at school ages, especially at 14-17 years (high school). Counties with remote reopening did not have increased incidence, which may also relate to their lower COVID-19 rate before the reopening, their public mask mandate or their social-economic advantage.

Remote learning consequences are not clear cut as there are tradeoffs, including increased inequities, less efficient learning and less monitoring for the growing risk of suicidality.^6,7^ Counties that reopened physically and had increased COVID-19 incidence could consider remote learning, especially at the high school level.

## Supporting information

STROBE checklist

## Data Availability

Data availability: publicly available from the Florida Department of Health at https://www.arcgis.com/home/item.html?id=4cc62b3a510949c7a8167f6baa3e069d

## ARTICLE INFORMATION

### Author Contributions

Oren Miron had full access to all of the data in the study and takes responsibility for the integrity of the data and the accuracy of the data analysis.

### Concept and design

All authors.

### Acquisition, analysis, or interpretation of data

All authors.

### Drafting of the manuscript

Oren Miron.

### Critical revision of the manuscript for important intellectual content

Yu, Wilf-Miron, Kohane and Davidovitch.

### Statistical analysis

Oren Miron, Kohane and Yu.

### Study supervision

Kohane and Davidovitch.

### Data availability

publicly available from the Florida Department of Health at https://www.arcgis.com/home/item.html?id=4cc62b3a510949c7a8167f6baa3e069d

